# A blood biomarker of accelerated aging in the body associates with worse structural integrity in the brain: replication across three cohorts

**DOI:** 10.1101/2023.09.06.23295140

**Authors:** Ethan T. Whitman, Calen P. Ryan, Wickliffe C. Abraham, Angela Addae, David L. Corcoran, Maxwell L. Elliott, Sean Hogan, David Ireland, Ross Keenan, Annchen R. Knodt, Tracy R. Melzer, Richie Poulton, Sandhya Ramrakha, Karen Sugden, Benjamin S. Williams, Jiayi Zhou, Ahmad R. Hariri, Daniel W. Belsky, Terrie E. Moffitt, Avshalom Caspi, the Alzheimer’s Disease Neuroimaging Initiative

## Abstract

Biological aging is the correlated decline of multi-organ system integrity central to the etiology of many age-related diseases. A novel epigenetic measure of biological aging, DunedinPACE, is associated with cognitive dysfunction, incident dementia, and mortality. Here, we tested for associations between DunedinPACE and structural MRI phenotypes in three datasets spanning midlife to advanced age: the Dunedin Study (age=45 years), the Framingham Heart Study Offspring Cohort (mean age=63 years), and the Alzheimer’s Disease Neuroimaging Initiative (mean age=75 years). We also tested four additional epigenetic measures of aging: the Horvath clock, the Hannum clock, PhenoAge, and GrimAge. Across all datasets (total N observations=3,380; total N individuals=2,322), faster DunedinPACE was associated with lower total brain volume, lower hippocampal volume, and thinner cortex. In two datasets, faster DunedinPACE was associated with greater burden of white matter hyperintensities. Across all measures, DunedinPACE and GrimAge had the strongest and most consistent associations with brain phenotypes. Our findings suggest that single timepoint measures of multi-organ decline such as DunedinPACE could be useful for gauging nervous system health.

## INTRODUCTION

Aging is the primary risk factor for many prevalent diseases (Niccoli and Partridge, 2012). Indeed, geroscientists have begun to treat aging itself as a preventable cause of many aging-related diseases (Barzilai et al., 2018; Campisi et al., 2019; Matt and Rabinovitch, 2015). The *geroscience hypothesis* defines aging as the gradual, progressive, correlated biological decline of the entire body over decades (Kennedy et al., 2014; Vadim, 2016). Crucially, individuals of the same *chronological* age often vary in their rate of *biological* aging (Belsky et al., 2015). Despite its importance, there is still no agreed-upon measure of biological aging (Ferrucci et al., 2020). To address this, researchers have begun to use DNA methylation to quantify aging. DNA methylation is a highly age-sensitive epigenetic process wherein methyl groups are selectively added to DNA molecules to affect gene transcription. Attempts to develop measures of aging have often used blood DNA methylation because blood is the most widely profiled source of DNA and blood DNA methylation is a biological substrate that is sensitive to age-related changes across the body (Horvath and Raj, 2018; Levine et al., 2015).

In the past decade, several algorithms have been developed to estimate biological aging using DNA methylation (Rutledge et al., 2022). These algorithms are typically referred to as ‘epigenetic clocks.’ The first generation of epigenetic clocks were trained largely on chronological age (Hannum et al., 2013; Horvath, 2013). Subsequently, a second generation of clocks was trained on cross-sectional measures of current health that predict mortality (Levine et al., 2018; Lu et al., 2019). Tests for associations between first– and second-generation epigenetic clocks and structural brain integrity have yielded mixed results. Some studies reported large positive associations between accelerated epigenetic age and poorer structural brain integrity (Davis et al., 2017; Hillary et al., 2021; Proskovec et al., 2020), while others reported small positive associations (Milicic et al., 2022; Raina et al., 2017), and still others reported null associations or even associations in the opposite direction (Cheong et al., 2022; Chouliaras et al., 2018).

In contrast to these earlier epigenetic clocks, we recently developed a third-generation DNA methylation-based measure that is unique in estimating a person’s rate of biological aging. The DunedinPACE (**P**ace of **A**ging **C**alculated from the **E**pigenome) algorithm was developed by first measuring people’s rate of physiological change over time and then identifying the methylation patterns that optimally captured individual differences in their age-related decline (Belsky et al., 2022). Specifically, age-related decline was measured over ages 26, 32, 38, and 45 years in 19 biomarkers of the cardiovascular, metabolic, renal, immune, dental, and pulmonary organ systems among healthy midlife individuals of the same chronological age participating in the Dunedin Study (Elliott et al., 2021b). Methylation patterns at the end of the 20-year observation period were then identified that estimated how fast each participants’ multi-organ decline occurred during the 20 years leading up to the point of measurement (Belsky et al., 2022). Thus, Dunedin PACE was designed to capture methylation patterns reflecting individual differences in the rate of age-related multi-organ decline and it has been robustly associated with multimorbidity and mortality (Belsky et al., 2022; Bernabeu et al., 2023; Faul et al., 2023; Kuiper et al., 2023; Lachlan et al., n.d.; McMurran et al., 2023). Importantly, Dunedin PACE allows for readily measuring the pace of aging in individuals who lack data to implement longitudinal physiological profiling.

Of note, Dunedin PACE was not trained on any measures of central nervous system decline. Thus, it is not clear whether Dunedin PACE is associated with structural brain integrity. Recent studies have used longitudinal multi-organ measurements to demonstrate that aging ‘below the neck’ is related to aging of the brain (Elliott et al., 2021a, 2021b; Tian et al., 2023); however, it is unknown whether a measurement from a blood sample at a single timepoint also captures the association between brain and body aging. Dunedin PACE has been associated with cognitive and clinical measures that are thought to index health of the central nervous system. For instance, faster Dunedin PACE has been associated with more rapid cognitive decline (Belsky et al., 2022; Reed et al., 2022), mild cognitive impairment, and dementia (Sugden et al., 2022). These findings suggest that Dunedin PACE indexes typical decline of cognitive ability during typical aging as well as in neurodegenerative illness. Associations between DunedinPACE and these key cognitive and clinical phenotypes suggest that DunedinPACE may also be associated with structural brain integrity, though this question has not been formally tested.

We examined associations between DunedinPACE and multiple measures of brain structure across three large datasets spanning mid-to late-life: the Dunedin Study (N=770, mean age=45 years), the Framingham Heart Study Offspring Cohort (FHS-OC; N=903, mean age=63.76 years), and the Alzheimer’s Disease Neuroimaging Initiative (ADNI; N observations=1,707, N individuals=649; mean age=75.41 years). Across all three datasets (N observations=3,380; N individuals=2,322), we tested for associations between DunedinPACE and measures of structural brain integrity derived from high-resolution magnetic resonance imaging (MRI; **Figure 1**) including: total brain volume (TBV), hippocampal grey matter volume (HC), white matter hyperintensity (WMH), mean cortical thickness (CT), and total cortical surface area (SA). In addition, we leveraged the longitudinal nature of ADNI to test for associations between DunedinPACE and age-related changes in structural brain integrity. For comparison, we also tested associations between first– and second-generation epigenetic clocks and structural brain integrity.

**Figure 1.**
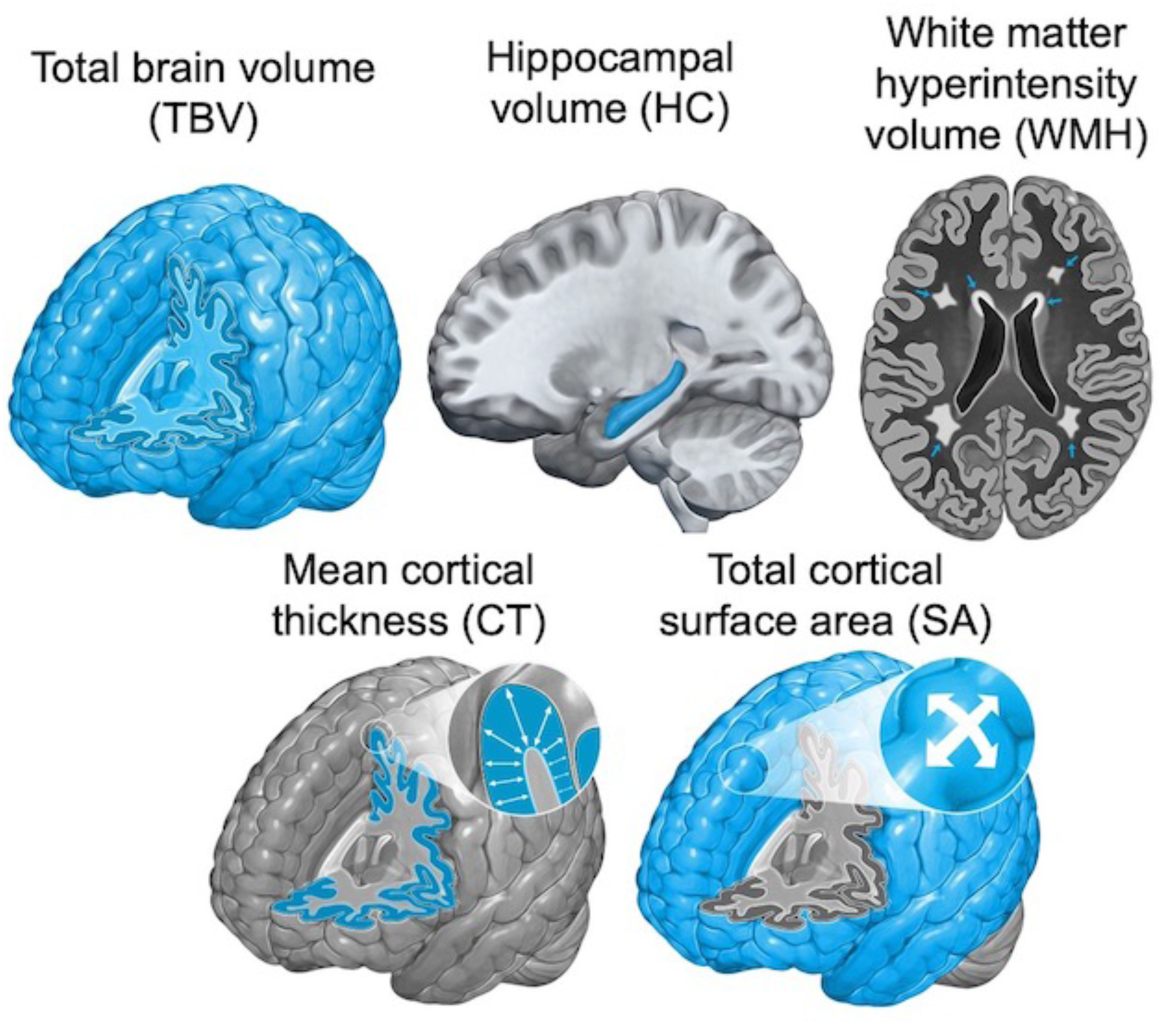
Diagrams of MRI-derived brain structural integrity measures. Phenotype of interest is represented in blue. Top row shows total brain volume (TBV), hippocampal volume (HC), and white matter hyperintensity volume (WMH). Bottom row shows mean cortical thickness (CT) and total cortical surface area (SA).

## METHODS

Data used in the current analyses were collected in the Dunedin Study (Poulton et al., 2023), the FHS-OC (Feinleib et al., 1975), and ADNI (Petersen et al., 2010). Further details on each of these studies is provided below. All analyses were checked for reproducibility by an independent data analyst who used the manuscript to derive code and reproduce statistics in an independent copy of the data.

### Dunedin Study

The Dunedin Study is a longitudinal study of a population-representative birth cohort born between April 1972 and March 1973 in Dunedin, New Zealand (Poulton et al., 2023). Assessments were carried out at birth and ages 3, 5, 7, 9, 11, 13, 15, 18, 21, 26, 32, 38, and most recently 45 years, when 94% of living members took part. DNA methylation and MRI data reported here were collected at the age-45 assessment phase. The Dunedin Study was approved by the New Zealand Health and Disability Ethics Committee and the Duke University Institutional Review Board. All Study members provided written informed consent.

#### DNA Methylation

DNA methylation was measured from whole blood using Illumina Infinium MethylationEPIC BeadChip Arrays and run at the Molecular Genomics Shared Resource at the Duke Molecular Physiology Institute. Details on DNA methylation methods in the Dunedin Study have been reported previously (Belsky et al., 2022).

#### MRI

T1-weighted and T2-weighted fluid attenuated inversion recovery (FLAIR) images were collected using a Magnetom Skyra 3T scanner. TBV, SA, CT, and HC were generated from the T1-weighted images using The Human Connectome Project minimal preprocessing pipeline and WMH from the T2-weighted images using the Unidentified Bright Object detector with visual confirmation of accuracy provided by a trained neuroradiologist (Glasser et al., 2013; Jiang et al., 2018). Further details on MRI methods in the Dunedin Study have been reported previously (Arbeloff et al., 2019).

### Framingham Heart Study – Offspring Cohort (FHS-OC)

The FHS tracks the development of cardiovascular disease in three generations of families from Framingham, Massachusetts, beginning in 1948 (Tsao and Vasan, 2015). We analyzed data collected from the second generation of participants, known as the Offspring Cohort (Feinleib et al., 1975). All DNA methylation data were collected at the Framingham Offspring 8^th^ follow-up, and MRI data were collected during an independent schedule of assessments conducted approximate to the time of the 8^th^ follow-up. We obtained Offspring Cohort data for this project from the National Institutes of Health Database of Genotypes and Phenotypes (dbGaP) under accession phs000007.v33.p14. DNA methylation data are available as substudy phs000724.v10.p14. Brain imaging data are available as substudy phs002559.v1.p14. We analyzed DNA methylation data downloaded as phg000492.v5.FHS_DNAMethylation.raw-data-idat and brain imaging data downloaded as phs000007.v33.pht004364.v2.t_mrbrfs_2010_1_0900s. The FHS was approved by the Institutional Review Board for Human Research at Boston University Medical Center. All participants provided written informed consent.

#### DNA Methylation

DNA methylation was measured from whole blood using Illumina Infinium HumanMethylation450 BeadChip Arrays and run at the University of Minnesota and The Johns Hopkins University (dbGaP phs000724.v9.p13). Further details of DNA methylation methods in the FHS-OC have been reported previously (Mendelson et al., 2017).

#### MRI

T1-weighted images were collected using a Siemens Magnetom 1.5T scanner. TBV, SA, CT, and HC were generated using FreeSurfer version 5.3 (Fischl, 2012). As T2-weighted FLAIR images were not available in FHS-OC, we used FreeSurfer estimates of white matter hypointensity volume derived from T1-weighted images. We consider white matter hypointensity volume in FHS-OC collectively alongside white matter hyperintensity volume in the Dunedin Study and ADNI, as both measures index white matter lesions, are highly correlated with one another, and show similar associations with age and Alzheimer’s disease pathology (Wei et al., 2019). Further details on MRI methods in FHS-OC have been reported previously (McGrath et al., 2019; Van Lent et al., 2023).

### Alzheimer’s Disease Neuroimaging Initiative (ADNI)

The primary goal of ADNI is to test whether serial magnetic resonance imaging (MRI), positron emission tomography (PET), other biological markers, and clinical and neuropsychological assessments can be combined to measure the progression of neurodegeneration in individuals with mild cognitive impairment, Alzheimer’s disease, and healthy older adults. DNA methylation data were downloaded from the ADNI data repository on June 3^rd^, 2020 and MRI data were downloaded on June 12^th^, 2022 (adni.loni.usc.edu). ADNI was approved by the Institutional Review Boards of all the participating institutions. All participants provided written informed consent.

#### DNA Methylation

DNA methylation was measured from whole blood using the Illumina Infinium MethylationEPIC BeadChip Array and run at AbbVie. Further details on DNA methylation methods in ADNI have been reported previously (Sugden et al., 2022).

#### MRI

T1-weighted and T2-weighted FLAIR images were collected using either 1.5T or 3T scanners. TBV, SA, CT, and HC were generated from the T1-weighted images using FreeSurfer version 4.3, 5.1, or 6.0 (Fischl, 2012). WMH were generated from the T2-weighted images using a previously described algorithm (Decarli et al., 1999). MRI acquisition parameters varied across ADNI sites and waves (Jack et al., 2008); however, all MRI data underwent centralized quality control by ADNI investigators prior to becoming available for download. ADNI CT and SA data are distributed according to the FreeSurfer version used in processing; therefore, we used Longitudinal ComBat to harmonize across FreeSurver versions using all FreeSurfer observations with QC ratings of ‘Pass’ from individuals with DNA methylation data (Beer et al., 2020). Further details on MRI methods in ADNI can be found at adni.loni.usc.edu.

### DunedinPACE estimation

In all three datasets, we derived DunedinPACE from the DNA methylation data using the publicly available algorithm (https://github.com/danbelsky/DunedinPACE). Following prior work (Sugden et al., 2022), we residualized DunedinPACE for chronological age in the mixed-age FHS-OC and ADNI datasets. Because all Study members were aged 45 at the time of their blood draw and MRI scans, this step was not necessary for analyses of Dunedin Study data. Within each study, DunedinPACE values were standardized to mean=0, standard deviation=1. Further details of estimating DunedinPACE in the Dunedin Study, FHS-OC, and ADNI have been reported previously (Belsky et al., 2022; Sugden et al., 2022).

To allow for comparison of DunedinPACE with other epigenetic clocks, we also report cross-sectional results for the Horvath, Hannum, GrimAge, and PhenoAge clocks (Hannum et al., 2013; Horvath, 2013; Levine et al., 2018; Lu et al., 2019). Additional details about the calculation of these epigenetic clocks can be found in the **supplemental materials.**

### Primary analyses

We first conducted a regression analysis of linear associations between DunedinPACE and each MRI measurement in all three datasets. Diagrams of each MRI measurements are presented in **Figure 1**. All regression models controlled for sex. Models using FHS-OC and ADNI data also included covariates for age, age^2^, sex*age, and age^2^*sex to account for the age variation in these cohorts. We did not control for age in the Dunedin Study because all individuals were aged 45 at the time of blood draws and MRI. We also controlled for intracranial volume (ICV) for all analyses of TBV and HC. WMH volumes were log-transformed for normality prior to analyses. In analyses using ADNI data, we included multiple timepoints within individuals in our models and calculated robust standard errors to account for non-independence.

We leveraged the longitudinal nature of ADNI to test whether baseline DunedinPACE could predict the rate of subsequent decline in structural brain integrity. We restricted this analysis to data from a subset of ADNI participants who remained cognitively normal throughout enrollment (N=153). This is because neurodegeneration in Alzheimer’s disease may diverge from the normative aging patterns indexed by DunedinPACE. Next, we identified participants from this subset who had ≥3 timepoints for each MRI measure after their first DNA methylation timepoint (N=107-147, **Table 2**). Using this subset of cognitively normal participants with sufficient longitudinal MRI data, we generated multilevel linear models for each MRI measure with random effects for both person and age. Using these models, we derived trajectories to track decline in each MRI measure for each person (i.e., a “decline” curve). To focus on relative changes in structural brain integrity, we residualized TBV and HC for ICV prior to generating these curves. We then tested whether each person’s initial DunedinPACE measure could predict their subsequent rate of structural brain integrity decline, after controlling for age, sex, and length of observation period.

### Sensitivity analyses

We conducted two sensitivity analyses. First, white blood cell abundance is thought to affect whole-blood epigenetic clock estimates. Therefore, we repeated our analyses while controlling for white blood cell abundance (specifically plasmablasts, +CD8pCD28nCD45RA-T cells, naïve CD8 T cells, CD4 T cells, Natural Killer cells, monocytes, and granulocytes) estimated from DNA methylation (Horvath, 2013; Houseman et al., 2012). Second, carriership of the *APOE* ε4 risk-allele has been associated with altered structural brain integrity (Régy et al., 2022). Therefore, we repeated all analyses while including risk-allele carriership as a covariate to test whether *APOE* ε4 could account for any observed associations between DunedinPACE and structural brain integrity.

## RESULTS

### Demographic characteristics

In the Dunedin Study, 770 Study members (male=51.0%) had both DNA methylation and MRI data available at age 45. Details showing that these Study members continue to represent the original birth cohort can be found in the **supplemental materials.** Of note, 8 Dunedin Study members were missing *APOE* ε4 genotype data and were excluded from sensitivity analyses controlling for *APOE* ε4 carriership. In FHS-OC, 903 (male=42%) participants had both DNA methylation and MRI data. The mean age at the time of blood sample collection was 63.76 years (SD=8.11 years) and the mean age at the time of MRI was 64.53±8.14 years. Of note, only 820 participants in FHS-OC had *APOE* ε4 genotype data. Thus, sensitivity analyses controlling for *APOE* ε4 carriership in FHS-OC were performed on a smaller subsample than the main analyses. In ADNI, 649 participants (male=55.6%) had DNA methylation data: 83 had only a baseline measurement, 121 had two measurements, 407 had three measurements, 29 had four measurements, and 9 had five measurements. This yielded 1,707 overall DNA methylation samples. The mean age at blood sample collection was 75.41±7.66 years. After pairing DNA methylation timepoints with unique MRI observations that took place at a similar time (i.e., <6 months), there was some variation in the total number of observations and individuals included for each MRI measure. Descriptions of the subsamples for each measure in ADNI are presented in **Table 1**. Age distributions for all three datasets are shown in **Figure 2**.

**Figure 2.**
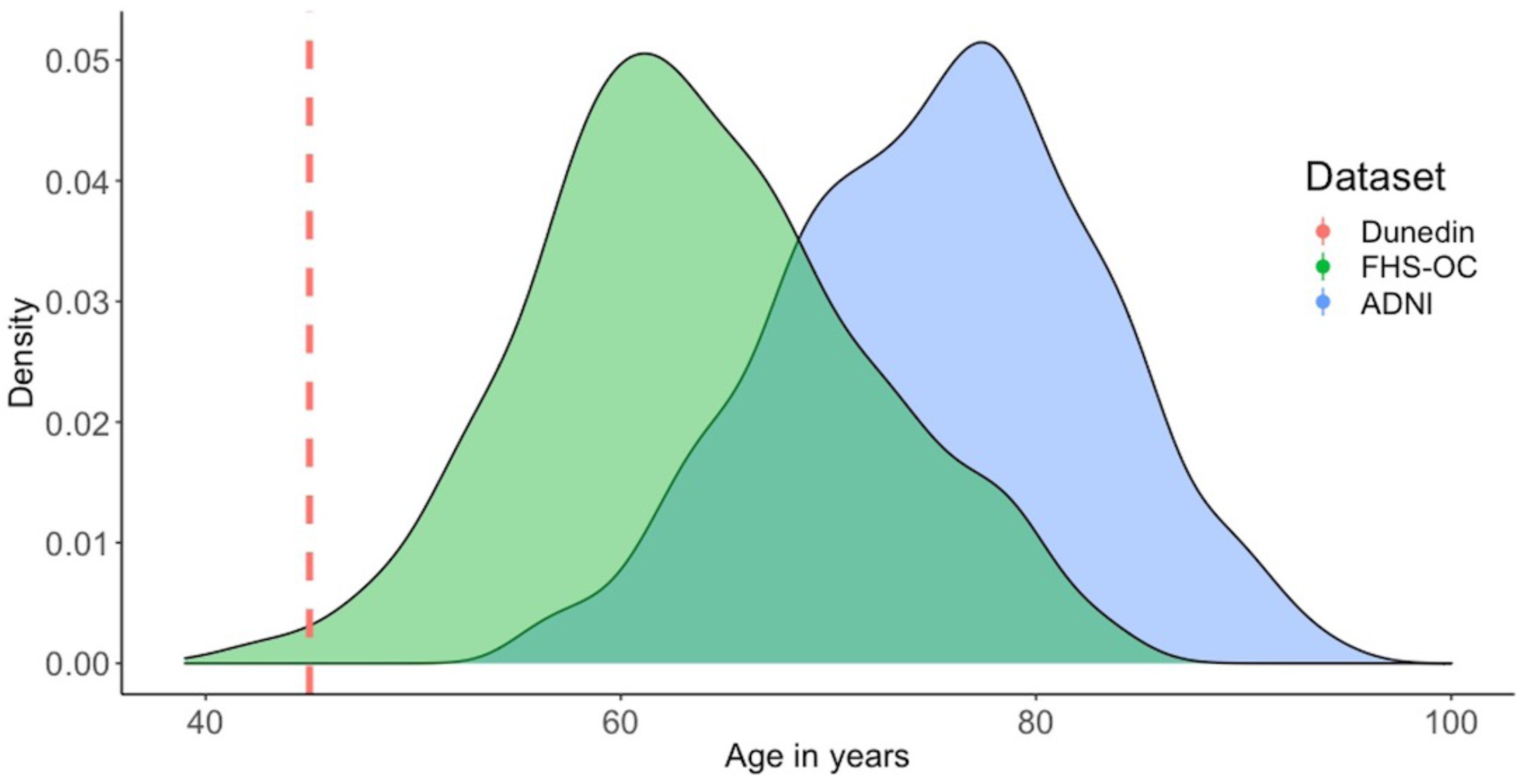
Age distributions in the Dunedin Study, FHS-OC, and ADNI. Density plots showing the age distributions in years at the time of DNA methylation observation in each dataset. Abbreviations: ADNI = Alzheimer’s Disease Neuroimaging Initiative; FHS-OC: Framingham Heart Study – Offspring Cohort

**Table 1.** Demographic information for the ADNI DNA methylation sample. Leftmost columns show the sample size for paired observations from ADNI. We have included both the total number of unique observations as well as the number of individuals. The third from right column presents the mean and standard deviations of the age in years at which the blood draw occurred. The fourth from the right column shows the time in days between the blood draw and MRI. Rightmost three columns show the proportion of most recent diagnostic status (cognitively normal, mild cognitive impairment, or dementia) at the time of the blood draw.

|  | **N** |  | **Mean Age in Years**<br>**at Collection (SD)** | **Mean days between MRI**<br>**Scan and DNA Methylation** | **CN (%)** | **MCI (%)** | **Dementia (%)** |
| --- | --- | --- | --- | --- | --- | --- | --- |
|  | Observations | Individuals |  |  |  |  |  |
| **DNA methylation** | 1,707 | 649 (55.6% male) | 75.4 (7.66) | - | 31.90% | 48.10% | 19.90% |
| Overlap with: |  |  |  |  |  |  |  |
| **TBV** | 1,426 | 604 (55.0% male) | 74.6 (7.50) | 2.0 | 31.9% | 50.9% | 17.2% |
| **HC** | 1,358 | 581 (54.9% male) | 74.4 (7.43) | 3.6 | 31.9% | 51.2% | 16.9% |
| **WMH** | 1,241 | 487 (54.0% male) | 73.6 (7.40) | 9.5 | 29.1% | 55.4% | 15.5% |
| **CT** | 1,078 | 446 (51.8% male) | 74.4 (7.37) | 10.1 | 31.6% | 50.6% | 17.7% |
| **SA** | 1,078 | 446 (51.8% male) | 74.4 (7.37) | 10.1 | 31.6% | 50.6% | 17.7% |

Abbreviations: ADNI = Alzheimer's Disease Neuroimaging Initiative; CN = cognitively normal; MCI = mild cognitive impairment; TBV = total brain volume; HC = hippocampal volume; WMH = white matter hyperintensities; CT = mean cortical thickness; SA = total cortical surface area

### Total brain volume

Across all three datasets, people with accelerated biological aging, as indexed by faster DunedinPACE, had lower TBV (Dunedin Study: β=–.06, p<.001, 95% CI: [–.09, –.03]; FHS-OC: β=–.04, p=.03, 95% CI: [–.07, –.004]; ADNI: β=–.06, p = .005, 95% CI: [–.10, –.02]; **Figure 3**). These associations were robust to white blood cell abundance and APOE ε4 carriership (see **supplemental materials**).

**Figure 3.**
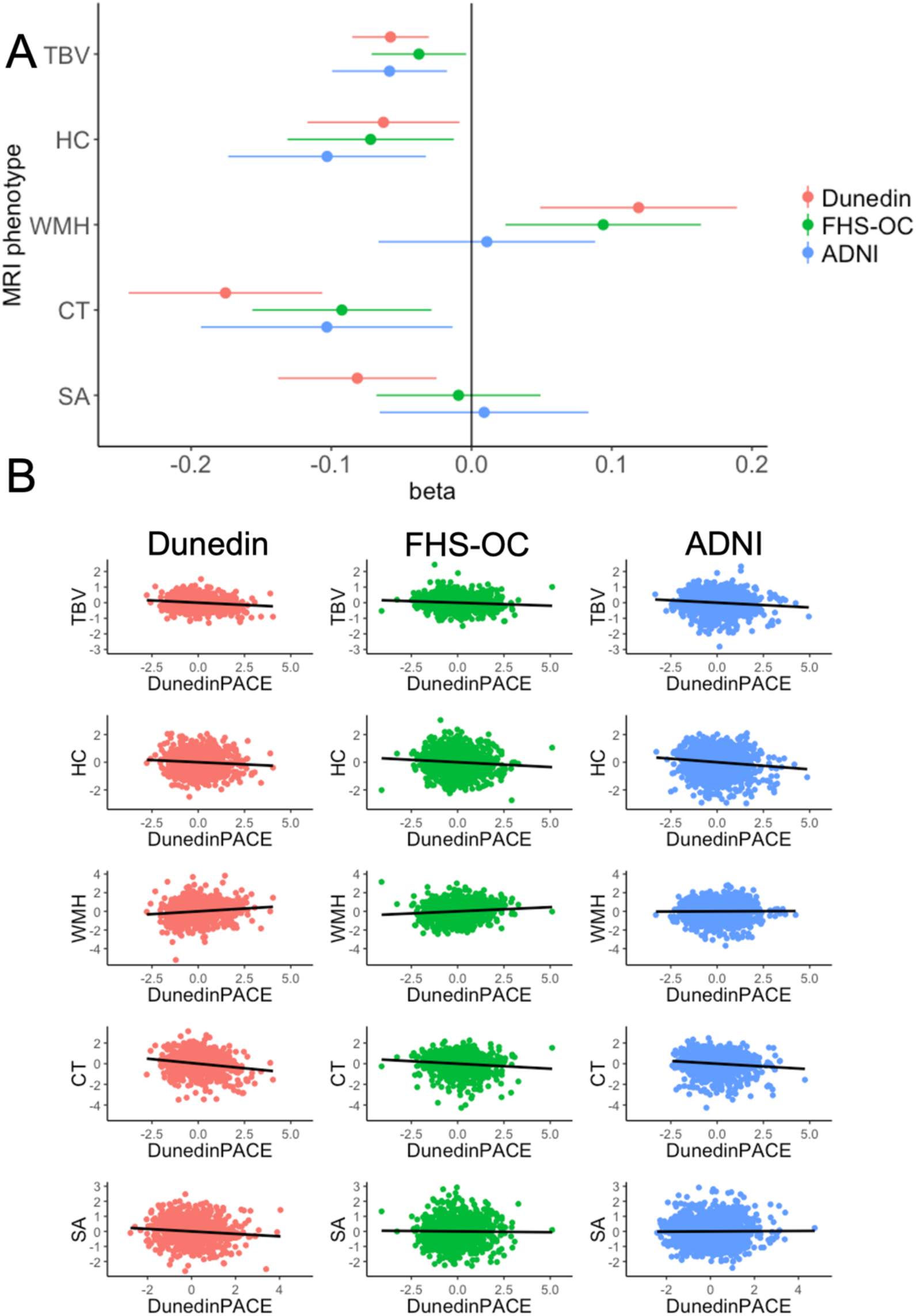
Associations between DunedinPACE and structural brain integrity. DunedinPACE was negatively associated with TBV, CT, and HC across all three datasets, and positively associated with WMH volume in the Dunedin Study and FHS-OC. A) Forest plot of all associations reported as standardized betas (error bars represent 95% confidence intervals). B) Scatterplots of all associations. X-axes represent scaled MRI measures. Y-axes represent scaled values for DunedinPACE. Abbreviations: ADNI = Alzheimer’s Disease Neuroimaging Initiative; FHS-OC: Framingham Heart Study – Offspring Cohort; TBV = total brain volume; HC = hippocampal grey matter volume; WMH = white matter hyperintensities volume, CT = mean cortical thickness, SA = total cortical surface area.

### Hippocampal grey matter volume

Across all three datasets, people with faster DunedinPACE had lower HC (Dunedin Study: β=–.06, p=.02, 95% CI: [–.12, –.01]; FHS-OC: β=–.07, p=.02, 95% CI: [–.13, –.01]; ADNI: β=–.10, p=.004, 95% CI: [–.17, –.03]; **Figure 3**). These associations were robust to white blood cell abundance and APOE ε4 carriership (see supplemental materials).

### White matter hyperintensities

People with faster DunedinPACE had greater WMH volume in the Dunedin Study (β=.12, p=.001, 95% CI: [–.19, –.05]; **Figure 3**) and the FHS-OC (β=.09, p=.01, 95% CI: [.02, .16]; **Figure 3**). We did not observe a significant association between DunedinPACE and WMH volume in ADNI (β=.01, p=.78, 95% CI: [–.07, .09]; **Figure 3**). These results were similar when controlling for white blood cell abundance or APOE ε4 carriership (see **supplemental materials**).

### Cortical thickness

Across all three datasets, people with faster DunedinPACE had thinner cerebral cortex (Dunedin Study: β=–.18, p<.001, 95% CI: [–.24, –.11]; FHS-OC: β=–.09, p=.01, 95% CI: [–.16, –.03]; ADNI: β=–.10, p=.02, 95% CI: [–.19, –.01]; **Figure 3**). These associations were robust to white blood cell abundance and APOE ε4 carriership (see **supplemental materials**).

### Cortical surface area

In the Dunedin Study, people who were aging faster, as measured by DunedinPACE, tended to have less total cortical surface area (β=–.08, p=.01, 95% CI: [–.14, –.02], **Figure 3**). However, we did not observe this association in FHS-OC or ADNI (FHS-OC: β=–.01, p=.75, 95% CI: [–.07, .05]; ADNI: β=.01, p=.81, 95% CI: [–.07, .08]; **Figure 3**). These results were not affected by white blood cell abundance or APOE ε4 allele carriership (see **supplemental materials**).

### Change in structural brain integrity

In ADNI, 153 participants with DNA methylation data remained cognitively normal throughout enrollment. Of these, varying numbers had MRI measures at a minimum of three timepoints allowing for calculation of change trajectories (**Table 2**). Analyses of data from these participants revealed expected age-related changes in all MRI measures (see **supplemental materials**). We did not observe significant associations between baseline DunedinPACE and subsequent change in MRI phenotypes (TBV: β=–.01, p=.92, 95% CI: [–.18, .16]; HC: β=–.01, p=.95, 95% CI: [–.18, .17]; WMH: (β=–.08, p=.38, 95% CI: [–.26, .10]; CT: β=–.06, p=.57, 95% CI: [–.27, .15]; SA: β=-19, p=.06, 95% CI: [–.38, .004]).

**Table 2.**
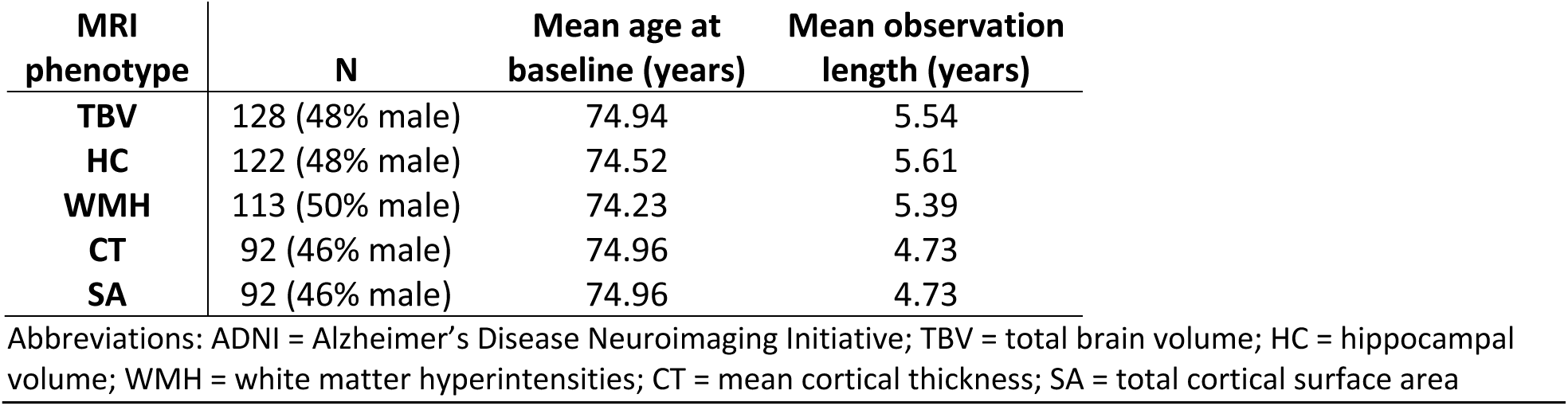
Demographic description for the ADNI longitudinal MRI sample. Sample size, sex, average age, and average observation length for ADNI participants who remained cognitively normal during ADNI enrollment and with ≥ 3 MRI timepoints after DNA methylation observation.

### Comparison with other epigenetic clocks

For comparison purposes, we repeated our cross-sectional analyses replacing DunedinPACE with each of four well-studied epigenetic clocks: first-generation Horvath and Hannnum clocks, and second-generation GrimAge and PhenoAge clocks. Associations of first-generation clocks with MRI measures were mostly null. PhenoAge generally had non-significant associations that were smaller than DunedinPACE. GrimAge had significant associations of similar magnitude to DunedinPACE in the Dunedin Study and FHS-OC; however, GrimAge only had a significant association with CT in ADNI. Complete details of these analyses are presented in **supplemental materials.**

## DISCUSSION

By aggregating three large datasets, we assembled the largest sample to date for examining associations between epigenetic clocks and brain anatomy. Using this sample, we showed that DunedinPACE, an epigenetic index of an individual’s pace of biological aging derived by tracking multi-organ decline from age 26 to 45 in a same-age birth cohort of healthy people is associated with several MRI measures of structural brain integrity in three independent datasets encompassing mid-to late-life. In all three datasets, people with accelerated biological aging, as indexed by faster DunedinPACE, had smaller total brain volume, smaller hippocampal grey matter volume, and thinner average cortex. In two datasets, people with faster DunedinPACE also had greater burden of white matter hyperintensities, a clinical MRI measure of white matter lesions. These results support the geroscience hypothesis of correlated decline across the entire body (Kennedy et al., 2014) by showing that faster decline of non-nervous organ systems (i.e., cardiovascular, metabolic, renal, immune, dental, and pulmonary) is associated with worse structural brain integrity during midlife and older age. Our findings align with recent work showing that aging of body organ systems is associated with aging of the brain (Elliott et al., 2021a; Tian et al., 2023), and further demonstrate that this association can be captured using a blood sample from a single timepoint.

The associations between DunedinPACE and MRI measures of structural brain integrity were consistent across our three datasets despite differences in cohort demographics. The Dunedin Study, FHS-OC, and ADNI differ substantially in participant age (Dunedin Study: age=45; FHS-OC mean age=64; ADNI mean age=75; **Figure 2**). ADNI is also highly enriched for Alzheimer’s disease and related dementias, whereas the Dunedin Study and FHS-OC are not. Approximately 18% of the ADNI participants in our analyses had been diagnosed with Alzheimer’s disease at the time of assessment. In contrast, nobody in either the Dunedin Study or FHS-OC datasets was diagnosed with dementia. Notably, associations between DunedinPACE and structural brain integrity across all three datasets are similar in magnitude to those between direct, longitudinal measures of organ-system decline and MRI measures in the Dunedin Study (Elliott et al., 2021b). Thus, a single timepoint of DunedinPACE closely replicates associations between brain and body aging estimated from longitudinal measurements of multiple organ systems. The fact that the observed effect sizes are generally consistent between the Dunedin Study, FHS-OC, and ADNI further suggests good generalizability of DunedinPACE across age and cognitive status.

Our findings indicate that DunedinPACE had strong and consistent associations with structural brain integrity relative to first-generation epigenetic clocks. Across all three datasets used here, associations between first-generation epigenetic clocks and brain structure were null or very close to zero. Previous neuroimaging research using first-generation epigenetic clocks has yielded inconsistent associations with brain structure (Cheong et al., 2022; Chouliaras et al., 2018; Davis et al., 2017; Proskovec et al., 2020; Raina et al., 2017). This is likely due to underpowered study designs (Liu et al., 2023). While we find that first-generation clocks tend to show null associations with MRI measures, a second-generation clock, GrimAge, was associated with brain integrity in both Dunedin and FHS-OC, although, GrimAge associations with TBV and HC were null in ADNI. Overall, our findings suggest that newer epigenetic clocks such as DunedinPACE and GrimAge may have greater promise for gauging structural brain integrity relative to first-generation epigenetic clocks. This is in line with the growing consensus that aging biomarkers trained only on chronological age have limited ability to detect age-related health outcomes whereas aging biomarkers trained on the *rate* of biological aging are more associated with to age-related health outcomes (Morqri et al., 2023; Zhou et al., 2022).

Associations between DunedinPACE and WMH in ADNI were null, in contrast to the significant associations observed in the Dunedin Study and FHS-OC. One potential explanation for this discrepancy is that ADNI is highly enriched for participants with high levels of amyloid-β, the accumulation of which can contribute to the formation of WMH in Alzheimer’s disease as well as cerebral amyloid angiopathy (Eider et al., 2019; Greenberg et al., 2020; Weaver et al., 2019). Thus, the pathophysiology of WMH in these conditions may not be as closely associated with the normative biological aging processes indexed by DunedinPACE, limiting the degree to which DunedinPACE may be associated with WMH in ADNI.

We did not find evidence that baseline DunedinPACE predicted subsequent risk-related change in brain structure in ADNI participants who remained cognitively normal throughout enrollment. However, the analytic sample with the requisite three MRI scans to model trajectories of reliable change in structural brain integrity was small. This reduced statistical power and limited our ability to detect associations. Nevertheless, our analyses provide circumstantial evidence that DunedinPACE is associated with neurodegenerative change. Specifically, in all three datasets, DunedinPACE was associated with TBV controlling for ICV, which represents the ‘maximal’ size a person’s brain reached during their life as the cranial cavity does not shrink during aging (Royle et al., 2013). Thus, “TBV controlling for ICV” can be interpreted as a proxy measure for longitudinal atrophic decline in TBV since childhood (Royle et al., 2013). While speculative, this suggests the hypothesis that faster DunedinPACE could be associated with accelerated neurodegeneration. Research using larger samples is needed to test this hypothesis.

Our findings are consistent with a recent report using data from the UKBiobank (Tian et al., 2023) to reveal that change in multiple individual organ systems (i.e., musculoskeletal, cardiac, metabolic, and pulmonary) each mapped onto change in the brain. Here we demonstrate a similar brain-body connection using DunedinPACE, a single timepoint DNA methylation measure trained on longitudinal decline in multiple organ systems. These two reports using complementary designs show that MRI measures of structural brain integrity are correlated with progressive, multi-organ decline in the rest of the body. Future studies should longitudinally measure multiple organ systems to address which is more relevant for brain aging: decline unique to individual organs, or the shared decline across organs predicted by the geroscience hypothesis and operationalized through DunedinPACE.

Our study has limitations. First, most of the data analyzed were derived from White participants reflecting the paucity of racial and ethnic diversity in datasets that have both DNA methylation and brain MRI. Notably, there is evidence that DunedinPACE can index health outcomes amongst both Black and Asian individuals (Graf et al., 2022; Schmitz et al., 2022; Schmitz and Duque, 2022; Shen et al., 2023). Second, DunedinPACE does not necessarily represent methylation of specific genes thought to contribute to neurodegeneration or aging progression, and it is best thought of as a non-causal statistical indicator of multi-organ decline. Finally, we were only able to assess change in structural brain integrity in one dataset, ADNI, which is highly enriched for Alzheimer’s disease, had the smallest number of individuals without dementia, and contained many of the oldest individuals (i.e., age>80 years). We recommend larger, more diverse, and more population-representative studies to assess how DunedinPACE is related to change in structural brain integrity.

We provide evidence that accelerated biological aging, as indexed by faster DunedinPACE, is consistently associated with structural brain integrity across three large datasets spanning midlife to older age. This suggests that the geroscience hypothesis of correlated, progressive decline of organs includes the central nervous system. We build on prior work by demonstrating that associations between brain and body aging can be detected using a single timepoint whole blood measure of DNA methylation. Collectively, these findings reinforce that aging is a whole-body process and suggest that aging neuroscience research stands to benefit from further attention to organs ‘below the neck.’

## Data Availability

Dunedin Study data are available via managed access (https://moffittcaspi.trinity.duke.edu/research).
Framingham Offspring Study data openly available openly from dbGaP (phs000007.v33.p14, phs000724.v10.p14, and phs002559.v1.p14).
ADNI data is openly available at https://adni.loni.usc.edu/.

## ACKNOWLEDGEMENTS

This research received support from US-National Institute on Aging grants R01AG069939, R01AG032282, and R01AG049789 and UK Medical Research Council grants MR/P005918/1, and MR/X021149/1. DWB received support as a Fellow of the Canadian Institute for Advanced Research Child Brain Development Network.

We thank the Dunedin Study members, Unit research staff, and Study founder Phil Silva. The Dunedin Multidisciplinary Health and Development Research Unit is supported by the New Zealand Health Research Council (Programme Grant 16-604) and New Zealand Ministry of Business, Innovation and Employment (MBIE). The Dunedin Unit is within the Ngāi Tahu tribal area who we acknowledge as first peoples, tangata whenua (translation: people of this land). We would like to acknowledge the assistance of the Duke Molecular Physiology Institute Molecular Genomics Core for their generation of data for the manuscript.

Data collection and sharing for this project was funded by the Alzheimer’s Disease Neuroimaging Initiative (ADNI) (National Institutes of Health Grant U01 AG024904) and DOD ADNI (Department of Defense award number W81XWH-12-2-0012). ADNI is funded by the National Institute on Aging, the National Institute of Biomedical Imaging and Bioengineering, and through generous contributions from the following: AbbVie, Alzheimer’s Association; Alzheimer’s Drug Discovery Foundation; Araclon Biotech; BioClinica, Inc.; Biogen; Bristol-Myers Squibb Company; CereSpir, Inc.; Cogstate; Eisai Inc.; Elan Pharmaceuticals, Inc.; Eli Lilly and Company; EuroImmun; F. Hoffmann-La Roche Ltd and its affiliated company Genentech, Inc.; Fujirebio; GE Healthcare; IXICO Ltd.; Janssen Alzheimer Immunotherapy Research & Development, LLC.; Johnson & Johnson Pharmaceutical Research & Development LLC.; Lumosity; Lundbeck; Merck & Co., Inc.; Meso Scale Diagnostics, LLC.; NeuroRx Research; Neurotrack Technologies; Novartis Pharmaceuticals Corporation; Pfizer Inc.; Piramal Imaging; Servier; Takeda Pharmaceutical Company; and Transition Therapeutics. The Canadian Institutes of Health Research is providing funds to support ADNI clinical sites in Canada. Private sector contributions are facilitated by the Foundation for the National Institutes of Health (www.fnih.org). The grantee organization is the Northern California Institute for Research and Education, and the study is coordinated by the Alzheimer’s Therapeutic Research Institute at the University of Southern California. ADNI data are disseminated by the Laboratory for Neuro Imaging at the University of Southern California.

The Framingham Heart Study is conducted and supported by the National Heart, Lung, and Blood Institute (NHLBI) in collaboration with Boston University (Contract No. N01-HC-25195, HHSN268201500001I and 75N92019D00031). Funding support for the Framingham Dementia Survival Information for All Cohorts dataset was provided by NIA grant R01-AG054076. This manuscript was not prepared in collaboration with investigators of the Framingham Heart Study and does not necessarily reflect the opinions or views of the Framingham Heart Study, Boston University, or NHLBI.

## DISCLOSURE STATEMENT

K. Sugden, A. Caspi, D. W. Belsky, D. L. Corcoran, R. Poulton, and T. E. Moffit are listed as inventors of DunedinPACE, a Duke University and University of Otago invention licensed to TruDiagnostic Inc. All other authors report no conflict of interest.

## Notes

### Author Declarations

The New Zealand Health and Disability Ethics Committee and the Duke University Institutional Review Board gave ethical approval for the Dunedin Study. This work used only openly available human data from the Framingham Offspring Study obtained from dbGaP (phs000007.v33.p14, phs000724.v10.p14, and phs002559.v1.p14). This work used only openly available human data from ADNI obtained from https://adni.loni.usc.edu/. ADNI was approved by the Institutional Review Boards of all participating institutions. Further information can be found at https://adni.loni.usc.edu/.

